# Unraveling the clinicopathological and molecular changes induced by neoadjuvant chemotherapy and endocrine therapy in hormone receptor-positive/HER2-low and HER2-0 breast cancer

**DOI:** 10.1101/2023.12.27.23299114

**Authors:** Francesco Schettini, Sabrina Nucera, Fara Brasó-Maristany, Irene De Santo, Tomás Pascual, Milana Bergamino, Patricia Galván, Benedetta Conte, Elia Seguí, Isabel García Fructuoso, Raquel Gómez Bravo, Pablo Rivera, Ana Belén Rodríguez, Olga Martínez-Sáez, Sergi Ganau, Esther Sanfeliu, Blanca González-Farre, Maria Vidal, Barbara Adamo, Isaac Cebrecos, Eduard Mension, Gabriela Oses, Pedro Jares, Sergi Vidal-Sicart, Meritxell Mollà, Montserrat Muñoz, Aleix Prat

**Affiliations:** Translational Genomics and Targeted Therapies in Solid Tumors, August Pi I Sunyer Biomedical Research Institute (IDIBAPS), Barcelona, Spain; Medical Oncology Department, Hospital Clinic of Barcelona, Barcelona, Spain; Faculty of Medicine and Health Sciences, University of Barcelona, Barcelona, Spain; Department of Human Pathology "G. Barresi", University of Messina, Messina, Italy; Medical Oncology Unit, Ave Gratia Plena Hospital, _San Felice a Cancello (CE)_, Italy; SOLTI Cooperative Research Group, Barcelona, Spain; Medical Oncology Department, Catalan Institute of Oncology, _Badalona_, Spain; Department of Radiology, Diagnosis Imaging Center, Hospital Clinic of Barcelona, Barcelona, Spain; Department of Pathology, Biomedical Diagnostic Center, Hospital Clinic of Barcelona, Barcelona, Spain; Department of Obstetrics and Gynecology, Hospital Clinic of Barcelona, Barcelona, Spain; Department of Nuclear Medicine, Diagnosis Imaging Center, Hospital Clinic of Barcelona, Barcelona, Spain; Radiation Oncology Department, Hospital Clínic de Barcelona, Barcelona, Spain; Institute of Cancer and Hematologic Diseases, Hospital Clinic of Barcelona, Barcelona, Spain; Breast Cancer Unit, Institute of Oncology Barcelona (IOB) – Quirónsalud, Barcelona, Spain; Reveal Genomic, Barcelona, Spain

**Keywords:** HER2-low, neoadjuvant chemotherapy, neoadjuvant endocrine therapy, hormone receptor-positive, HER2 dynamics

## Abstract

**Background:** The characterization and comparison of gene expression (GE) and intrinsic subtypes (IS) changes induced by neoadjuvant chemotherapy (NACT) and endocrine therapy (NET) in hormone receptor-positive(HR+)/HER2-low vs. HR+/HER2-0 breast cancer (BC) has not been conducted so far. Most evidence on the association of HER2 status with pathologic responses and prognosis in HR+/HER2-negative BC is controversial and restricted to NACT-treated disease. Similarly, a temporal heterogeneity in HER2 status has been described only with NACT.

**Methods:** We retrospectively recruited a consecutive cohort of 186 patients with stage I-IIIB HR+/HER2-negative BC treated with neoadjuvant therapy (NAT). Available diagnostic biopsies and surgical samples were characterized for main pathological features, PAM50 intrinsic subtypes (IS) and risk-of-relapse (ROR)-P score, and GE. Associations with pathologic complete response (pCR), residual cancer burden (RCB)-0/I, event-free survival (EFS) and overall survival (OS) based on HER2 status were assessed. Pre/post pathologic/molecular changes were analyzed in matched samples.

**Results:** The HER2-low (62.9%) and HER2-0 (37.1%) cohorts did not differ significantly in main baseline features, treatments administered, breast conserving surgery (BCS), pCR and RCB-0/I rates, EFS and OS. NAT induced, regardless of HER2 status, a significant reduction of ER/PgR and Ki67, a downregulation of PAM50 proliferation- and luminal-related genes/signatures, an upregulation of selected immune genes and a shift towards less aggressive IS and lower ROR-P. Moreover, 25% of HER2-0 changed to HER2-low and 34% HER2-low became HER2-0. HER2 shifts were significant after NACT (p<0.001), not NET (p=0.063), with consistent *ERBB2* mRNA level dynamics. HER2 changes were not associated to EFS/OS.

**Conclusions:** HER2 status changes after NAT in ∼1/4 of cases, mostly after NACT. Targeted adjuvant strategies should be investigated accordingly. Molecular downstaging with current chemo/endocrine agents and immunotherapy should not rely on HER2 immunohistochemical levels in HR+/HER2-negative BC. Instead, HER2-low-targeted approaches should be explored to pursue more effective and/or less toxic dimensional downstaging.

**Highlights:**

1. Hormone receptor-positive (HR+)/HER2-low and HER2-0 breast cancer (BC) showed similar post-neoadjuvant surgical outcomes.
2. Neoadjuvant therapy (NAT) induced a shift towards less aggressive subtypes and ROR-P classes regardless of HER2 status.
3. All NAT strategies induced a downregulation of proliferation- and luminal biology-related genes, regardless of HER2 status.
4. NAT induced changes in HER2 status, with a discordance rate of 34% and HER2-low showing higher instability than HER2-0.
5. HER2 status at baseline, after surgery and its dynamics were not significantly associated to long-term outcomes.

## Introduction

Breast cancer represents a heterogeneous disease. Molecularly, four intrinsic subtypes (IS) have been identified, namely Luminal A, Luminal B, HER2-enriched (HER2-E) and Basal-like, along with a Normal-like group^1^. For practical purposes, in order to broadly guide therapeutic choices in the clinic, a surrogate of these molecular subtypes is traditionally detected by immunohistochemistry (IHC), though with no perfect overlap^2^. Estrogen-dependent, Luminal A/B non-HER2 overexpressing tumors are usually regrouped into an hormone receptor-positive (HR+)/HER2-negative subset^3^. Nevertheless, non-luminal IS represent up to 10-30% of HR+/HER2-negative tumors^4^. In recent years, a novel subgroup of interest has emerged within the HER2-negative subset, following impressive therapeutic responses obtained to novel potent antibody-drug conjugates (ADCs) directed against HER2, especially trastuzumab-deruxtecan (T-DXd)^5,6^. This subgroup, currently named HER2-low, is characterized by low levels of expression of HER2, consisting in a IHC score of 1+, or 2+ with no amplification of the HER2 gene (*ERBB2*) by situ hybridization (ISH) techniques, according to the American Society of Clinical Oncologists (ASCO)/College of American Pathologists (CAP) guidelines^5^. Among HR+/HER2-negative breast cancer, HER2-low represents ∼65% of cases, while the rest has a HER2 IHC score of 0 (HER2-0)^7^.

In early-stage HR+/HER2-negative disease, neoadjuvant endocrine therapy (NET) or neoadjuvant chemotherapy (NACT) represent a therapeutic standard for inoperable disease to determine a tumor dimensional downstaging and achieve surgical resection. At the same time, the neoadjuvant approach might be useful in upfront operable disease to increase breast-conserving surgery (BCS) rates, to provide predictive and prognostically valuable clinicopathological and/or molecular information, to adapt further therapeutic strategies, either in the clinic or in the context of clinical trials^8,9^. Furthermore, the achievement of pathological complete response (pCR) or limited residual cancer burden (RCB) after NACT has been associated to better prognosis^10,11^. Nonetheless, the evidence in patients receiving NET is scarce. Data regarding the correlation between pathologic responses and prognosis in the HR+/HER2-low population are controversial and the evidence is mostly restricted to NACT-treated disease^12^. Moreover, a temporal heterogeneity in HER2 status has been described with NACT in mixed HR+ and HR-negative populations, with controversial impact on prognosis^13–15^. Finally, to our knowledge, a comprehensive biologic characterization in terms of gene expression (GE) and molecular subtypes changes induced by neoadjuvant treatments in HR+/HER2-low vs. HER2-0 disease has not been carried out so far.

We conducted Neoendo, a retrospective observational study to unveil the clinicopathological and molecular changes induced by NACT and NET broadly in HR+/HER2-negative breast cancer. Preliminary results were previously presented^16^. We hereby present a sub-analysis focused on assessing molecular and pathological changes induced by NACT and NET in HR+/HER2-low vs. HR+/HER2-0 breast cancer, in terms of PAM50 IS and risk-of-relapse score switch, gene expression changes, HER2 dynamic changes and potential prognostic implications. The ultimate purpose was to understand if HER2 status might play a role in the evolving neoadjuvant therapeutic landscape of HR+/HER2-negative breast cancer.

## Materials and methods

### Study population

A consecutive cohort of patients with stage I-IIIB HR+/HER2-negative breast cancer treated as *per* standard-of-care NACT or NET at the Hospital Clinic of Barcelona (HCB) between 2014-2018 was included in this study. NACT consisted of standard anthracycline and/or taxane-based regimens. NET consisted of 3 to 6 months of an aromatase inhibitor (AI) or tamoxifen. Neoadjuvant therapy information, surgical outcomes, and baseline tumor pathological features (i.e. at least HR levels, Ki67% and HER2 IHC score) had to be available for study inclusion. The electronic medical records were retrospectively reviewed to obtain the relevant information. Study objectives/endpoints are detailed in **Supplementary methods**.

### Pathology and PAM50 GE analysis

Estrogen receptor (ER), progesterone receptor (PgR), Ki67, stromal tumor-infiltrating lymphocytes (TILs) and HER2 status were assessed according to international guidelines^7,17–20^. HER2-low were defined as previously mentioned^21^. RNA was purified from available archival formalin-fixed paraffin-embedded (FFPE) tumor tissues from pre-treatment baseline diagnostic biopsies and surgical specimens, as previously described^22^ and analyzed at the nCounter platform (NanoString Technologies, Seattle, WA, USA) (**Supplementary methods**). The research-based PAM50 predictor was used to assign each tumor to an IS or to the Normal-like group^23^. The PAM50 risk-of-relapse score based on subtype and proliferation (ROR-P) and the PAM50 proliferation, luminal, HER2 and basal signatures were assessed as elsewhere described^23^.

### Statistical analysis

Standard unpaired/paired non-parametric test were used to compare continuous and categorical variables between HER2-0 and HER2-low cases and pre/post-surgical variations. Survival analyses were conducted with the Kaplan-Meier method and log-rank test. Associations of clinicopathological and molecular features with event-free survival (EFS) and overall survival (OS) were assessed with Cox regression models to estimate hazard ratios (HRs) with their 95% confidence intervals (CIs). Logistic regressions were conducted to estimate odds ratios (ORs) and 95% CIs of the associations with BCS, RCB and pCR. Significance was set at p≤0.05, without corrections for multiplicity due to exploratory purposes. Significance analysis of microarrays (SAM) was used to assess GE differences. A false discovery rate (FDR)≤5% was considered for significance (**Supplementary methods**). All analyses were conducted with R vers. 3.6.1^24^ and SPSS^®^ Statistics vers. 24 (IBM^®^, Armonk, NYC, USA) for MacOSX.

## Results

### Demographics

One-hundred-eighty-six patients undergoing neoadjuvant therapy were included; 37.1% had HER2-0 and 62.9% had HER2-low breast cancer (70.9% HER2 1+ and 29.1% HER2 2+/ISH negative) (**Supplementary figure 1**). We did not identify significant differences between the two groups in baseline clinicopathological features, except for a higher prevalence of lobular tumors (21.7% vs. 6.0%, p=0.019), a lower proportion of ROR-P intermediate, and a higher proportion of ROR-P high in the HER2-0 group, as compared to HER2-low (48.9% and 28.9% vs. 65.4% and 10.3%, p=0.028). Detailed clinicopathological characteristics according to HER2 status are reported in **Table 1**.

**Table 1.** Population characteristics.

| Demographics |  | HER2-0 |  | HER2-Low |  | P |
| --- | --- | --- | --- | --- | --- | --- |
|  |  | N | % | N | % |  |
|  |  | 69 | 37.1 | 117 | 62.9 |  |
| Age (years) |  |  |  |  |  |  |
|  | Median | 55.1 | - | 58.8 | - | 0.244 |
|  | IQR | 47.5 - 66.3 | - | 50.3 - 66.8 | - |  |
| Menopausal status |  |  |  |  |  |  |
|  | Pre/peri | 19 | 27.5 | 24 | 20.5 | 0.272 |
|  | Post | 50 | 72.5 | 93 | 79.5 |  |
| ER (%) |  |  |  |  |  |  |
|  | Median |  | 90.0 | - | 90.0 | 0.060 |
|  | IQR |  | 80.0 - 90.0 | - | 90.0 - 95.0 |  |
| PgR (%) |  |  |  |  |  |  |
|  | Median | 50 | - | 60 | - | 0.914 |
|  | IQR | 5.0 - 90.0 | - | 5.0 - 80.0 | - |  |
| HR status |  |  |  |  |  |  |
|  | HR-low (ER and PgR≤10%) | 2 | 2.9 | 3 | 2.6 | 0.892 |
|  | HR+ (ER and PgR>10%) | 67 | 97.1 | 114 | 97.4 |  |
| Ki67 (%) |  |  |  |  |  |  |
|  | Median | 22 | - | 21 | - | 0.295 |
|  | IQR | 11.0 - 35.0 | - | 10.0 - 26.0 | - |  |
| Stromal TILs (%) |  |  |  |  |  |  |
|  | Median | 5 | - | 1 | - | 0.295 |
|  | IQR | 1.0 - 5.0 | - | 1.0 - 5.0 | - |  |
|  | Overall | 66 | 95.7 | 115 | 98.3 |  |
| Grading |  |  |  |  |  |  |
|  | 1 | 11 | 17.2 | 30 | 27.0 | 0.086 |
|  | 2 | 44 | 68.8 | 74 | 66.7 |  |
|  | 3 | 9 | 14.1 | 7 | 6.3 |  |
|  | Overall | 64 | 92.8 | 111 | 94.9 |  |
| Histotype |  |  |  |  |  |  |
|  | Ductal/No special type | 50 | 72.5 | 101 | 86.3 | 0.019 |
|  | Lobular | 15 | 21.7 | 7 | 6.0 |  |
|  | Mixed/Other | 4 | 5.8 | 9 | 7.7 |  |
| **cT** |  |  |  |  |  |  |
|  | 1 | 21 | 30.4 | 43 | 36.8 |  |
|  | 2 | 38 | 55.1 | 60 | 51.3 | 0.658 |
|  | 3 - 4 | 10 | 14.5 | 14 | 12.0 |  |
| **cN** |  |  |  |  |  |  |
|  | 0 | 45 | 65.2 | 84 | 71.8 |  |
|  | 1 | 20 | 29.0 | 29 | 24.8 | 0.568 |
|  | 2 - 3 | 4 | 5.8 | 4 | 3.4 |  |
| **TNM** |  |  |  |  |  |  |
|  | I | 21 | 30.4 | 41 | 35.0 |  |
|  | II | 41 | 59.4 | 64 | 54.7 | 0.799 |
|  | IIIA-B | 7 | 10.1 | 12 | 10.3 |  |
| **Baseline PAM50 IS** |  |  |  |  |  |  |
|  | Luminal A | 20 | 40.8 | 48 | 54.5 |  |
|  | Luminal B | 19 | 38.8 | 29 | 33.0 |  |
|  | HER2-enriched | 4 | 8.2 | 6 | 6.8 | 0.391 |
|  | Basal-like | 2 | 4.1 | 3 | 3.4 |  |
|  | Normal-like | 4 | 8.2 | 2 | 2.3 |  |
|  | *Overall* | 49 | 71.0 | 88 | 75.2 |  |
| **Baseline ROR-P score** |  |  |  |  |  |  |
|  | Median | 40.8 | - | 30.7 | - | 0.087 |
|  | IQR | 13.4 – 53.2 | - | 13.0 – 43.2 | - |  |
|  | Low Risk | 10 | 22.2 | 19 | 24.4 |  |
|  | Intermediate Risk | 22 | 48.9 | 51 | 65.4 | 0.028 |
|  | High Risk | 13 | 28.9 | 8 | 10.3 |  |
|  | *Overall* | 45 | 65.2 | 78 | 66.7 |  |

**Legend.** IQR: interquartile range; ROR: risk of relapse; IS: intrinsic subtypes; TILs: tumor-infiltrating lymphocytes; HR: hormone receptor; ER: estrogen receptor; PgR: progesterone receptor; cT: clinical tumor size; cN: clinical axillary lymph-node status. Significant p values are reported in italics.

Approximately half patients *per* cohort received NACT (58.0% in HER2 and 48.7% in HER2-low). Of patients receiving NET, 18.8% in the HER2-0 and 12.8% in the HER2-low cohort, received posterior adjuvant CT. Almost all patients underwent adjuvant ET, but nine patients refused further treatment after surgery. There was no difference in BCS (p=0.693), nor in surgical axillary management (p=0.687). Overall, no significant differences in the neoadjuvant treatment received between the HER2-0 and HER2-low cohorts were seen (**Supplementary table 1**).

### Treatment-induced changes in ER, PR, HER2, KI67 and TILs

We explored potential neoadjuvant treatment-induced changes of ER, PgR, TILs, Ki67 and HER2 status in matched pre/post-treatment samples. The neoadjuvant treatment induced in HER2-low and HER2-0, respectively, a significant reduction in ER (p<0.001 and p=0.004), PgR (p<0.001 both) and Ki67 (p<0.001 both) levels, but not in TILs (p=0.603 and p=0.569). Post-surgical ER (p=0.826), PgR (p=0.146), Ki67 (p=0.867) and TILs (p=0.253) levels did not differ significantly between the HER2-0 and HER2-low cohorts. A significant modification in HER2 status from baseline to surgery was observed in the overall study population. More specifically, a net increase in the proportion of HER2-0 tumors (from 36.6% to 47.7%), with a reduction in the proportion of HER2-low (from 63.4% to 49.0%) was observed, along with few cases (3.3%) shifting towards a HER2+ status with a 3+ IHC score (p=0.001). The agreement between pre- and post-neoadjuvant HER2 status was only fair (Cohen’s K: 0.34, 95%CI: 0.20 – 0.48). However, while tumors treated with NACT showed statistically significant HER2 status modifications (p=0.009) comparable to what observed in the entire cohort, tumors receiving NET showed only a non-significant increase in the proportion of HER2-0 tumors, without shifts towards HER2+ disease (p=0.063). Coherently, *ERBB2* mRNA levels were significantly reduced in the overall population and in patients treated with NACT, but not in patients receiving NET (**Figure 1**).

**Figure 1.**
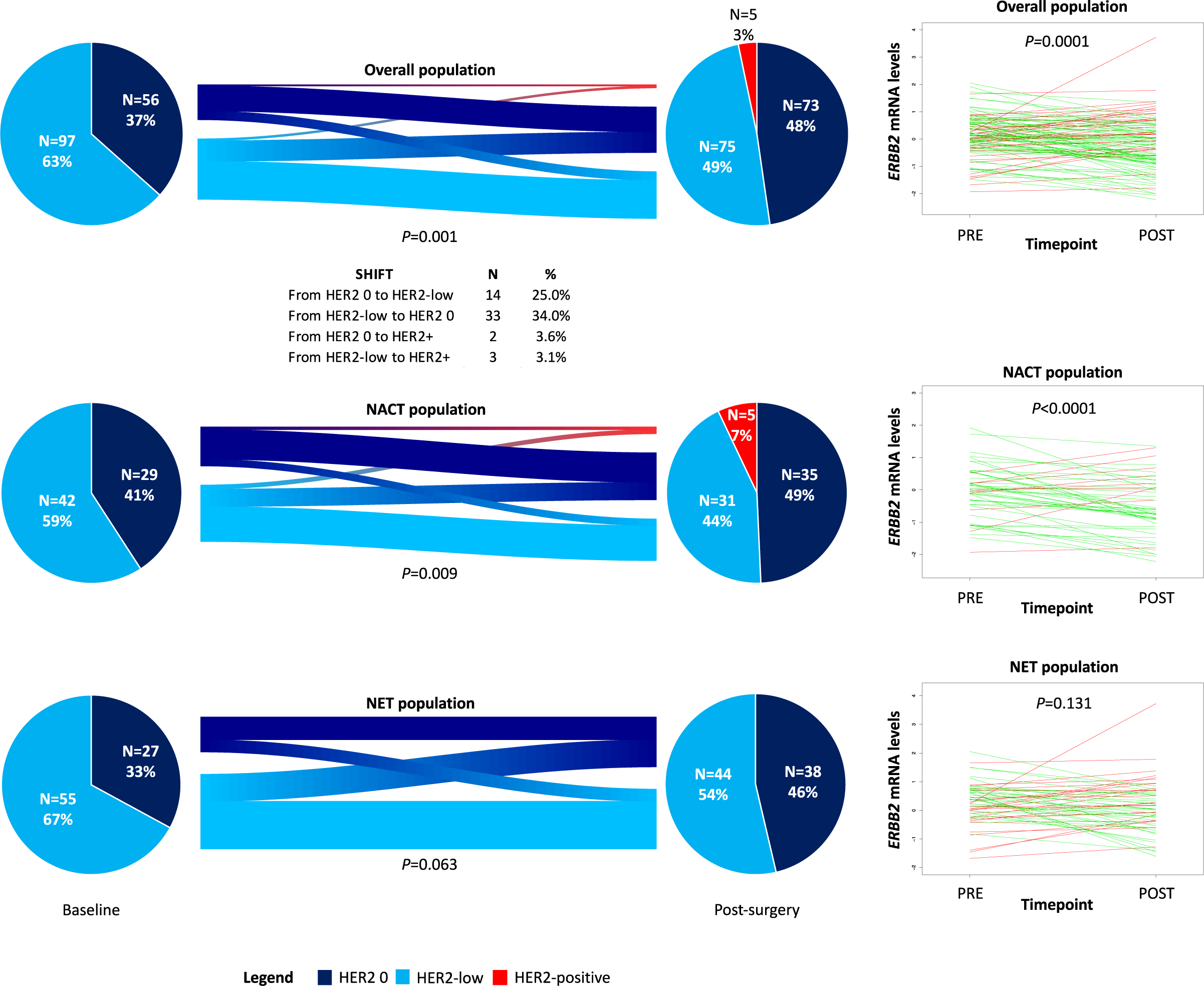
HER2 IHC and *ERBB2* mRNA levels changes from baseline to post-surgery in the overall population and according to neoadjuvant treatment. **Legend.** NAT: neoadjuvant therapy; NACT: neoadjuvant chemotherapy; NET: neoadjuvant endocrine therapy.

Baseline HER2 status was not significantly different between the NACT and NET cohorts (p=0.311), post-neoadjuvant status differed significantly (p=0.037) and the agreement between baseline and post-surgical HER2 status was only fair in both NET (Cohen’s K: 0.28, 95%CI: 0.07 – 0.48) and NACT (Cohen’s K: 0.40, 95%CI: 0.22 – 0.59) cohorts.

### Treatment-induced molecular changes

We explored potential neoadjuvant treatment-induced changes at the subtype, ROR, and genomic level in matched pre/post-treatment samples in the HER2-0 and HER2-low cohorts, separately, regardless of the neoadjuvant approach first, and then in separate NACT and NET cohorts. A significant switch towards the Luminal A IS and the Normal-like group was observed in both HER2-0 (p<0.001) and HER2-low (p<0.001) cohorts (**Figure 2A**). In accordance with unpaired samples, the distribution of PAM50 IS was similar between the two cohorts at baseline (p=0.384), but also after the neoadjuvant treatment (p=0.094) (**Supplementary table 2**). In both cohorts, a significant shift towards lower ROR-P categories was observed (**Figure 2B**). The ROR-P continuous score was significantly reduced from a median of 40.8 (interquartile range [IQR]: 13.1 – 55.0) to a median of −2.0 (IQR: −7.7 – 9.2) in the HER2-0 cohort (p<0.001). In the HER2-low cohort, ROR-P score was significantly reduced (p<0.001) from a median of 32.1 (IQR: 16.0 – 44.3) to −3.5 (IQR: −7.8 – 6.9) (**Supplementary table 2**). The distribution of ROR-P categories at baseline was slightly different between the two cohorts (p=0.034), consistently with what observed in unpaired samples (**Supplementary table 2**). After treatment, the proportion of ROR-P categories did not differ significantly between the two cohorts (p=0.272). ROR-P continuous score did not differ significantly at baseline (p=0.241) and after treatment (p=0.933) (**Supplementary table 2**).

**Figure 2.**
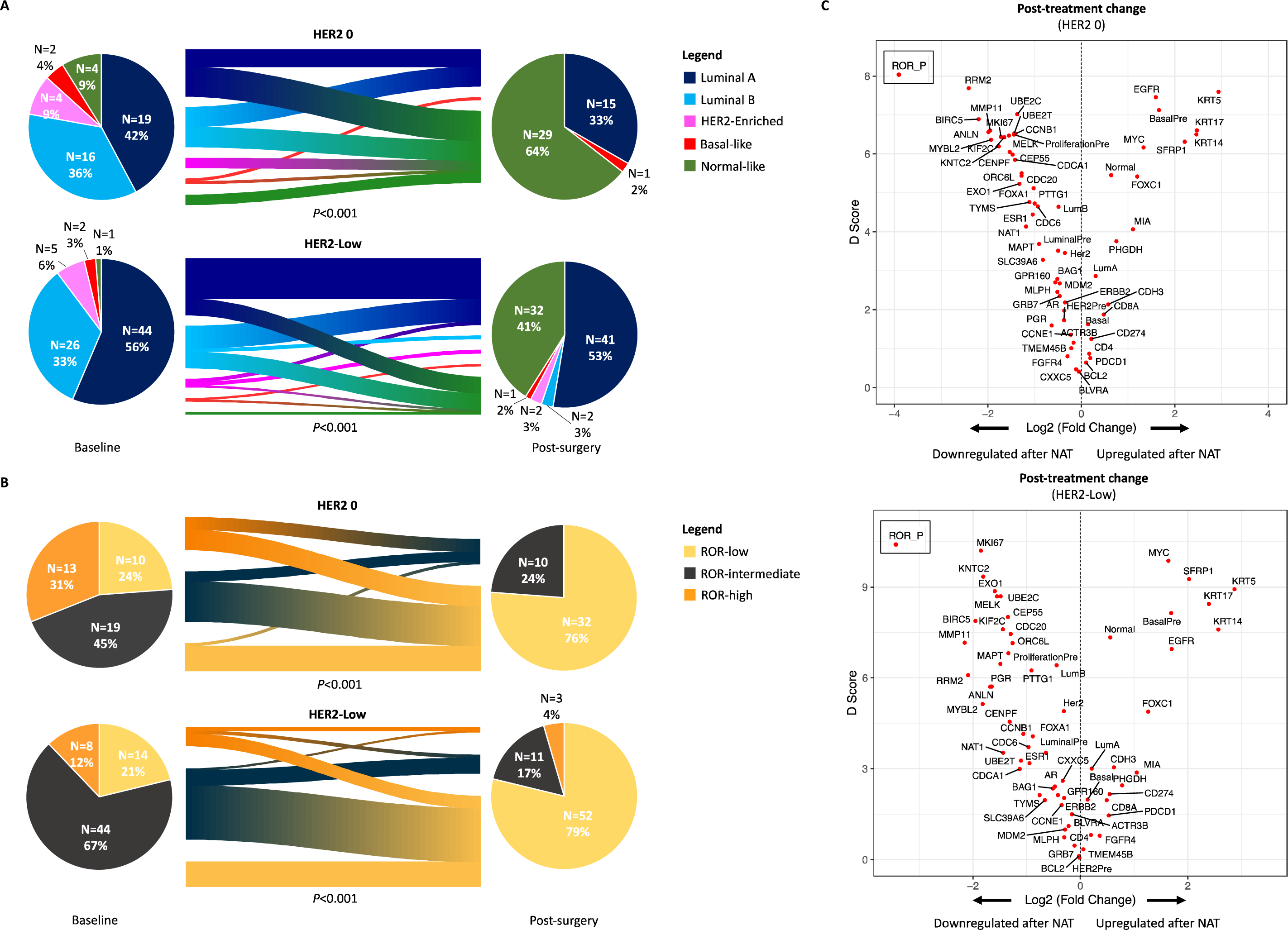
Treatment-induced molecular changes from baseline to post surgery according to HER2 status. **Legend. A:** PAM50 IS switch after neoadjuvant therapy according to HER2 status; **B:** PAM50 ROR-P class switch after neoadjuvant therapy according to HER2 status; **C:** gene expression changes after neoadjuvant therapy according to HER2 status. LuminalPre: luminal-related baseline genomic signature; HER2Pre: HER2 amplicon-related baseline genomic signature; BasalPre: basal-like-related baseline genomic signature; ProliferationPre: proliferation-related baseline genomic signatures; LumA: luminal A PAM50 intrinsic subtype correlation score; LumB: luminal B PAM50 intrinsic subtype correlation score; Her2: HER2-enriched PAM50 intrinsic subtype correlation score; Basal: basal-like PAM50 intrinsic subtype correlation score; Normal: normal-like PAM50 intrinsic subtype correlation score; ROR: risk of relapse score, here referred to research-based PAM50 ROR-P; D score: a T-statistic value that reflects a standardized change in expression. It measures the strength of the relationship between gene expression and the post-treatment category (versus pre-treatment); Red dots identify significantly differentially expressed genes for a false discovery rate (FDR)_≤_5%. The ROR-P dot originally fell outside of the established boundaries of the Volcano plots, due to a profound downregulation. To retain a good visualization of the plots, ROR-P dot has been separately added in the upper left quadrant without modifying the x-y axes’ scale.

When exploring NACT and NET effect, separately, within the HER2-0 cohort, we observed significant switches towards less aggressive PAM50 IS and lower risk ROR-P classes with both NACT (p<0.001 both) and NET (p<0.001 and p=0.009), consistent with the results of the overall cohort (**Supplementary table 3**). Similar findings were observed within the HER2-low cohort with NACT and NET in terms of PAM50 IS (p<0.001 both) and ROR-P class (p<0.001 both) switches (**Supplementary table 3**). No post-treatment difference in terms of PAM50 IS and ROR-P class distributions were observed between NACT and NET within the HER2-0 (p=0.499 and p=0.729, respectively) and HER2-low cohorts (p=0.100 and p=0.089, respectively). Similarly, neither significant differences were detected in terms of post-NACT PAM50 IS (p=0.499) and ROR-P classes (p=0.227) between HER2-0 and HER2-low, nor in post-NET PAM50 IS (p=0.100) and ROR-P classes (p=0.432).

At the GE level, a significant upregulation of basal-like-related genes/PAM50 signature and immune genes (i.e. *CD8A*, *PDCD1* and *CD274*) was observed after treatment, along with a significant downregulation of *CD4*, ROR-P score, luminal- and proliferation-related genes/PAM50 signatures (**Figure 2C**). When dissecting the results according to the neoadjuvant treatment adopted, we observed similar downregulation and upregulation patterns with NACT and NET in HER2-0 and HER2-low, separately (**Figure 3A**). In fact, most up/downregulated genes/signatures were shared between HER2-0 and HER2-low tumors, both with NACT (17 upregulated and 43 downregulated in common) and NET (16 upregulated and 41 downregulated in common) (**Figure 3B**). We then assessed baseline samples for potential upfront GE differences. HER2-low, in comparison to HER2-0, only showed a significant upregulation of the HER2 amplicon genes *ERBB2* and *GRB7* and luminal-related genes *NAT1* and *MMP11* (**Supplementary figure 2**). When comparing the two cohorts after neoadjuvant therapy (jointed NACT and NET), there were no significant GE differences, except for a persistent upregulation in the HER2-low cohort of the HER2 amplicon genes *ERBB2* and *GRB7*, associated to an upregulation of the PAM50 HER2 amplicon signature, as compared to HER2-0. Also, HER2-low tumors vs. HER2-0 showed an upregulation of the luminal-related genes *ESR1, MMP11* and *SLC39A6* (**Supplementary figure 2**). When separating the comparisons according to the neoadjuvant treatment received, only minimal differences with the jointed analysis were found (**Supplementary figure 2**).

**Figure 3.**
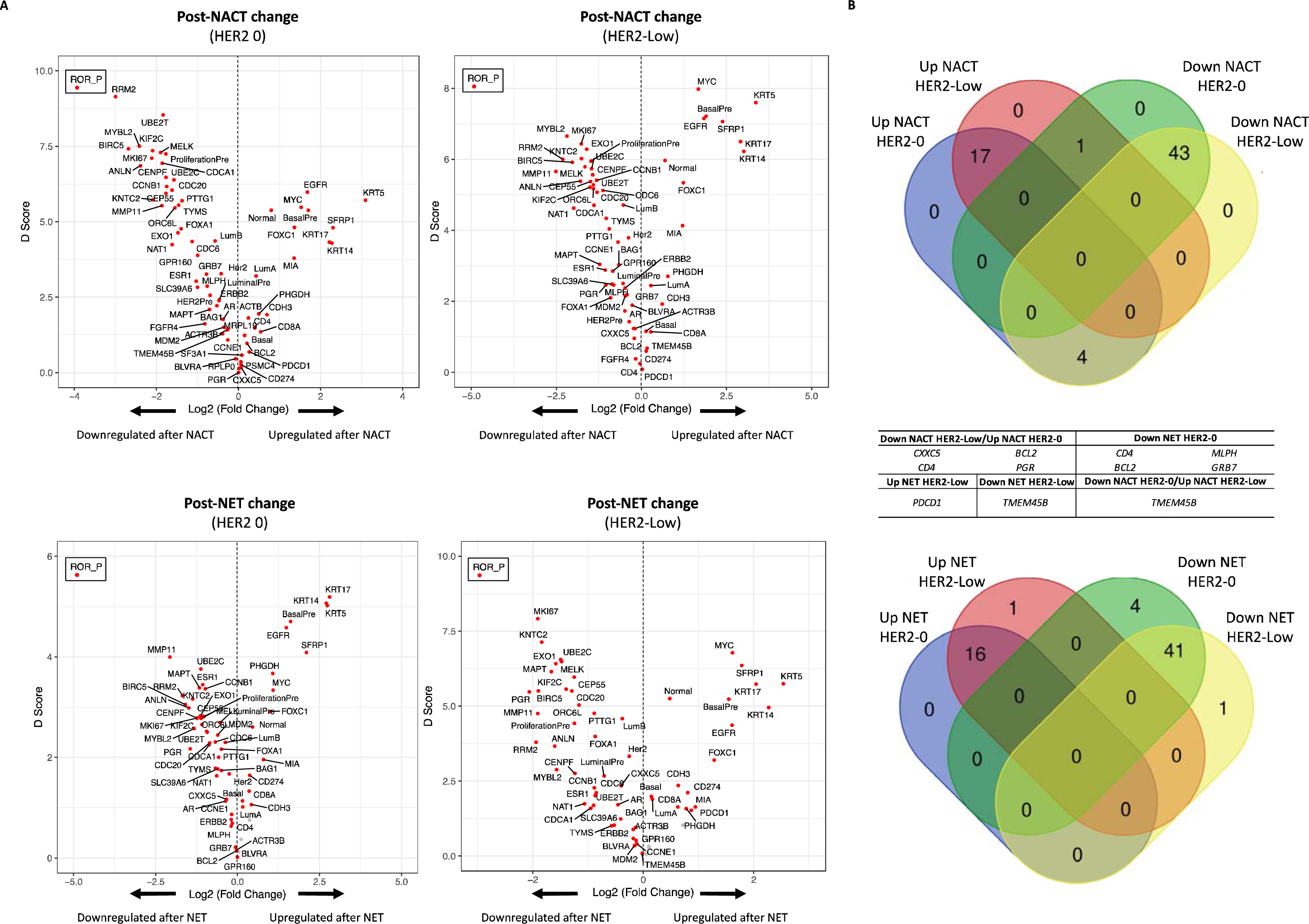
Gene expression changes induced by different neoadjuvant strategies according to HER2 status. **Legend. A:** Gene expression changes induced by NACT and NET in HER2-0 and HER2-low breast tumors; **B:** Venn diagrams of shared and uniquely up/downregulated genes between HER2-0 and HER2-low tumors, according to neoadjuvant treatment. Down: downregulated; Up: upregulated; NACT: neoadjuvant chemotherapy; NET: neoadjuvant endocrine therapy; LuminalPre: luminal-related baseline genomic signature; HER2Pre: HER2 amplicon-related baseline genomic signature; BasalPre: basal-like-related baseline genomic signature; ProliferationPre: proliferation-related baseline genomic signatures; LumA: luminal A PAM50 intrinsic subtype correlation score; LumB: luminal B PAM50 intrinsic subtype correlation score; Her2: HER2-enriched PAM50 intrinsic subtype correlation score; Basal: basal-like PAM50 intrinsic subtype correlation score; Normal: normal-like PAM50 intrinsic subtype correlation score; D score: a T-statistic value that reflects a standardized change in expression. It measures the strength of the relationship between gene expression and the post-treatment category (versus pre-treatment); Grey dots represent genes not differentially expressed, while red dots identify significantly differentially expressed genes for a false discovery rate (FDR)_≤_5%. The ROR-P dot originally fell outside of the established boundaries of the Volcano plots, due to a profound downregulation. To retain a good visualization of the plots, ROR-P dot has been separately added in the upper left quadrant without modifying the x-y axes’ scale.

### Pathological response and survival outcomes

A total of 11.3% of patients achieved a pCR/RCB-0, while 26.3% achieved RCB-0 or I. However, no significant differences in pCR and RCB-0/I rate were observed between HER2-0 and HER2-low cases (p=0.705 and p=0.740, respectively), as well as no significantly different HER2 status distribution according to pathology outcome (p=0.761). As such, HER2-low tumors, as compared to HER2-0, were not associated to better pathologic responses (**Figure 4A**). However, the pCR rate for patients receiving NACT was 18.6%, in comparison to the 3.4% pCR rate observed with NET. Patients had a significantly higher likelihood of achieving pCR (adjusted OR [OR_a_]: 4.48, p=0.030) and RCB-0/I (OR_a_: 4.00, p=0.001) than patients undergoing NET, irrespective of baseline Ki67, ER, PgR, HER2 status and TNM (**Supplementary table 4**). NACT, as compared to NET, was still independently more associated to pCR in the HER2-low cohort (OR_a_: 6.08, 95%CI: 1.15 – 32.30, p=0.034), and to RCB-0/I in both HER2-0 (OR_a_: 5.91, 95%CI: 1.13 – 30.91, p=0.035) and HER2-low (OR_a_: 3.36, 95%CI: 1.23 – 9.15, p=0.018) cohorts, irrespective of Ki67, ER, PgR and TNM.

**Figure 4.**
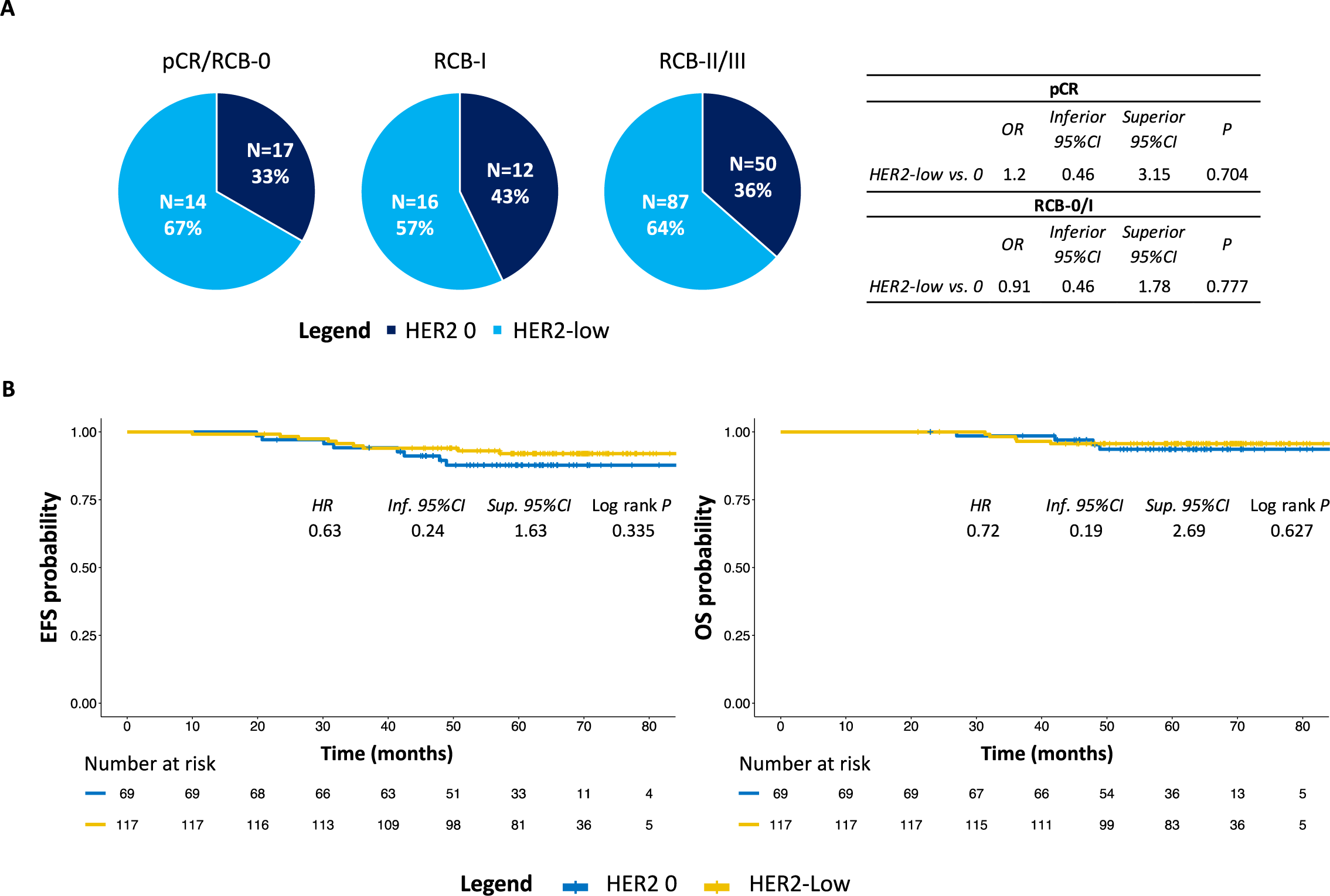
Association of HER2-0 and HER2-Low status with pathologic response and long-term outcomes. **Legend. A:** Pathologic outcomes according to HER2 status; **B:** EFS and OS according to HER2 status. Inf: inferior; Sup: superior; CI: confidence interval; OR: odds ratio; HR: hazard ratio; EFS: event-free survival; OS: overall survival; pCR: pathologic complete response; RCB: residual cancer burden.

At a median follow-up of 64.0 (95%CI: 62.4 – 66.7) months the median EFS and OS were not reached, with a total of 17 EFS events and 9 OS events occurred. Numerically lower 5-year EFS (87.7%, 95%CI: 80.1% - 96.1%) and OS (93.6%, 95%CI: 87.7 – 99.9%) were observed for the HER2-0 cohort, in comparison to the HER2-low cohort (5-year EFS 92.0%, 95%CI: 87.1 - 97.2%; 5-year OS 95.7%, 95%CI: 92.0 – 99.5%). However, no statistically significant differences were detected in terms of EFS (p=0.335) and OS (p=0.627), based on HER2 status at baseline (**Figure 4B**). Similarly, post-surgical HER2 status did not show an association with EFS (p=0.670) and OS (p=0.803). To note, at univariate analysis, a stability in HER2 IHC status from baseline to surgery was associated to worse EFS (p=0.003) and OS (p=0.030) than HER2 shifts (from 0 to low or positive and from low to 0 or positive). However, the result was not significant at the multivariable analyses for EFS (p=0.936) and OS (p=0.950) (**Supplementary table 5**). No specific baseline imbalances were observed between the HER2 stable and dynamic cohorts neither in the main tumor clinicopathological and molecular features (i.e. age, menopausal status, PgR, ER, TILs, Ki67, G, cT, cN, ROR-P, PAM50 IS), nor in the treatments received (all p>0.05; not shown). The only exception was represented by tumor histology, with a significantly higher proportion of lobular tumors (15.4% vs. 11.9%) in the group experiencing a HER2 shift, as compared to the group with sustained HER2 expression (p<0.001), which in turn showed a higher proportion of mixed tumors/tumors of rare histologies (8.9% vs. 3.8%).

## Discussion

As we and others had previously observed, HR+/HER2-low seems not to be a distinct subtype of breast cancer^7,25,26^. Nonetheless, while it shares with HR+/HER2-0 disease a similar tumor mutational burden and genomic landscape, HR+/HER2-low is characterized by a slightly higher proportion of luminal IS, a relatively lower expression of proliferation-related genes, along with a higher expression of *ERBB2* and luminal-related genes^7,25,26^. With these premises, we decided to investigate in a retrospective cohort from the HCB, the pathological and molecular effects induced by NACT and NET on HR+/HER2-low and HR+/HER2-0 disease, as well as the relationship of HER2 status and its dynamics, with surgical and long-term outcomes.

First, HER2-low predictive and prognostic role is controversial. Results are quite heterogeneous in the literature^27^. For this reason, a meta-analysis was recently conducted to shed light on this topic. Overall, in HR+/HER2-negative disease, HER2-low breast cancer was less associated with pCR, showed an association with slightly better OS and no differences in disease-free survival, in comparison to HER2-0^12^. In our study, HR+/HER2-low vs. HR+/HER2-0 disease showed only a limited 4.3% absolute benefit in EFS and 2.1% absolute benefit in OS at 5 years, without reaching statistical significance. Furthermore, patients with HER2-low and HER2-0 disease did not experience differential surgical outcomes in terms of BCS rates and pathologic responses. Importantly, NACT was confirmed to be associated to better pathologic responses than NET, independently of HER2 status and main baseline tumor features. Nevertheless, considering the prognostic improvements deriving from a significant tumor shrinkage after NACT^10,11^ and the limited pCR rates obtained with standard CT regimens in HR+ disease (usually ranging 10-20%^8^), confirmed in our cohort (∼19%), HR+/HER2-low tumors might gain benefit from alternative regimens including novel potent anti-HER2 ADCs, such as T-DXd. In this perspective, the currently ongoing TRIO-US B-12 TALENT is evaluating T-DXd, alone or in combination with the AI anastrozole as neoadjuvant treatment for HR+/HER2-low breast cancer^28^. Preliminary results showed a good activity for T-DXd in terms of objective responses, which translated in RCB-0/I rates of ∼15%, irrespective of AI addition, with surgical outcomes still pending for ∼1/4 of patients. These findings suggest that HER2-low-targeted strategies are still immature to enter the clinical practice scenario, but worthy exploring in the context of HR+/HER2-low early-stage breast cancer.

Noteworthy, both NACT and NET induced a significant reduction in ER, PgR and Ki67 levels in our cohort, regardless of HER2 status. The dynamics of these parameters, especially ER and Ki67, have been especially studied after NET and associated to long-term outcomes, allowing for possible tailoring of subsequent escalated or de-escalated therapeutic strategies according to patients’ prognosis^8^. Here, for the first time, we not only show that baseline HER2 status seems to have no impact on the dynamics of these IHC biomarkers, regardless of the neoadjuvant treatment administered, but also that these pathological changes were accompanied by consistent, similar modifications at the genomic level, in both HER2-low and HER2-0 tumors. We observed that the neoadjuvant therapy could induce a significant shift towards less aggressive tumor biology, represented here by a net increase in Normal-like group and Luminal A PAM50 IS, as well as a significant reduction in ROR-P score and risk class, accompanied by a downregulation in the PAM50 proliferation- and luminal biology-related genes/signature. These biologic effects were observed also when separating HER2-0 and HER2-low cases according to whether they had been treated with NACT or NET, with minimal differences based on treatment strategy. This molecular downstaging resembled what we observed in a previous analysis from the same population where NACT and NET were compared separately, regardless of HER2 status (i.e. mixed HER2-low and HER2-0 populations)^16^. In that same analysis, we observed an encouraging trend towards better survival outcomes associated to such a molecular downstaging^16^. Notably, Bergamino M. et al. observed a consistent molecular downstaging under AI presurgical treatment, as well^29^. A shift towards less aggressive subtypes and ROR score was also observed in the SOLTICORALEEN trial of patients with HR+/HER2-negative disease randomized to receive NACT or NET+CDK4/6-inhibitor ribociclib^30^. Since a new generation of clinical trials investigating molecular downstaging as a potential surrogate of better long-term outcomes and/or its role in sparing adjuvant CT in HR+/HER2-negative disease is underway (e.g. the RIBOLARIS trial^31^), our results highlight that no differential approaches should be envisioned specifically for HR+/HER2-low disease in this context.

Interestingly, Bergamino M. et al. also observed significant changes in the expression of immune response/immune-checkpoint components’ genes, that were associated with AI-resistant tumors^29^. In our cohort, although with a limited number of immune-related genes, namely *CD4, CD8A, PDCD1* (PD1) and *CD274* (PD-L1), we also observed a significant upregulation, with no differences between HER2-0 and HER2-low disease. These changes merit further investigation, especially after positive preliminary results from the KEYNOTE-756 trial, which showed that the addition of the anti-PD1 pembrolizumab to NACT in high risk HR+/HER2-negative breast tumors, can increase pCR rates^32^. Although differentiating between HER2-0 and HER2-low in this context does not seem to be critical, we cannot exclude that newer anti-HER2 immunoconjugates and bispecific antibodies might exert a differential immunomodulatory effect in HR+/HER2-low disease, as compared to HR+/HER2-0.

We and others previously demonstrated that the biology of HER2-low breast cancer is driven by HR status, is associated to a slightly higher prevalence of luminal IS and a higher levels of *ERBB2* mRNA levels than HER2-0 disease^7,25,33,34^. Our current results further confirm these findings, with HER2-low representing 63% of our cohort of HR+/HER2-negative breast tumors, accompanied by an overall similar baseline GE profile with HER2-0, but characterized by the upregulation of few HER2 amplicon and luminal-related genes, including *ERBB2*. Notably, after treatment, a significantly higher proportion of Luminal A IS in HER2-low than in HER2-0 disease was observed. This characteristic was associated to a significant upregulation of some luminal biology-related genes, including the crucial *ESR1*^1,35^, and a retained upregulation of *ERBB2*. Differences between NACT and NET were minimal in this regard, and with uncertain clinical relevance. However, neoadjuvant therapy induced a net reduction in *ERBB2* mRNA levels, both in HER2-0 and HER2-low disease. Changes in *ERBB2* mRNA levels were accompanied by HER2 status modifications at the IHC level. HER2 status discordance from baseline after neoadjuvant treatment was assessed in some recent studies focused on HER2-negative disease (including TNBC) undergoing NACT, and the discordance rates ranged between 26.4% - 53.8%^13–15,36^. In our analysis, focused on HR+ breast cancer, changes in HER2 status from the biopsy to the residual were observed, with a net increase in HER2-0 and a slight reduction of HER2-low tumors. The discordance rate was 34% overall, with HER2-low showing higher instability than HER2-0 (25% shifted from 0 to low and 34% from low to 0). However, the HER2 IHC switch seemed to be driven by the NACT subgroup, with a milder effect observed under NET. Similarly, the net reduction in *ERBB2* mRNA levels was significant in the overall population and in the subset of patients treated with NACT, but not in the NET cohort. These findings suggest that CT might have a more pronounced effect than ET on HER2 dynamics, although this should be confirmed in a larger cohort. Also, we cannot exclude that a baseline HER2 heterogeneity^37^ might have contributed, at least in part, to the observed HER2 status changes in residual disease.

In a study from Kang S. et al., the prognostic significance of HER2 dynamics was investigated, and no different long-term outcomes were observed according to HER2 changes in the HR+ subpopulation^13^. In our cohort, HER2 stability was accompanied by poorer long-term outcomes at univariate analysis, but the data was not confirmed when considering main prognostic clinicopathological factors, although a larger number of cases would be required to draw more appropriate conclusions. To note, there were no significant baseline differences, including in ROR-P class and PAM50 IS, except for a higher proportion of lobular disease in HER2 shifting cases. Still, histotypes were not prognostic in our cohort. Notably, 5 cases experienced a shift towards HER2+ disease, within both the HER2-0 (3.6%) and HER2-low (3.1%) disease, and 2 of these patients were treated with adjuvant anti-HER2-based therapy. Overall, the changes observed suggest that HER2 reassessment should be performed on residual disease, especially after NACT, with the purpose of exploring potential novel anti-HER2 directed post-neoadjuvant strategies in tumors shifting from 0 to low or positive disease. At present, a more practical reason to reassess HER2 status in residual disease would be also the need to detect an HER2-low disease so to grant access to T-DXd to patients with primary HER2-0 tumor, when re-biopsy is unfeasible in the metastatic context or when metastatic relapse is also HER2-0^21^.

Main study limitations were represented by the retrospective design and the relatively reduced number of cases. Moreover a 5-year median follow-up in a population with excellent long-term outcomes, limited our possibilities to detect statistically significant but clinically mild differences between HER2-low and HER2-0 cases. Another limitation consisted in the presence of missing pathological and genomic data, due to unavailability or scarce tumor cellularity issues. Nevertheless, >120 tumors were molecularly characterized and the number of 110 cases with pre/post paired samples for molecular analysis, with matched surgical and long-term outcomes is not easily achieved. In fact, none of the previous studies on the same topic provided an assessment of molecular treatment-induced GE, IS and molecular risk of relapse changes. Furthermore, to our knowledge, this is the first study focused specifically on HR+/HER2-low and HR+/HER2-0 undergoing neoadjuvant treatment, and is also the first study reporting HER2 status changes under NET and not only NACT. Additionally, it is the first time that changes at the protein level, as detected by IHC, were associated to the study of *ERBB2* mRNA dynamics, supporting the concept that HER2 shifts are not only a methodological artifact or the results of inter-pathologist disagreement in HER2 assessment. Finally, despite the absence of randomization, the HER2-0 and HER2-low cohorts at baseline were sufficiently well balanced with respect to main clinicopathological and genomic features considered.

To conclude, NACT and NET modify tumor biology of HR+/HER-negative breast tumors, regardless of HER2 status. The clinicopathological and molecular changes induced by NACT and NET in HR+/HER2-low and HER2-0 disease seem not to differ significantly, suggesting that current approaches focused on molecular downstaging and immunotherapy should not rely on HER2 status assessment in this context. However, the role of new agents like potent anti-HER2 ADCs is still unknown. Moreover, novel HER2-low-targeted approaches should be further investigated to pursue a more effective and/or less toxic dimensional downstaging in HR+/HER2-low disease, than what is currently obtained with standard approaches. Finally, HER2 status change occurs in a significant proportion of cases, especially after NACT, with consistent *ERBB2* mRNA levels’ modifications, supporting the need of HER2 status reassessment in the post-neoadjuvant scenario.

## Ethic statement

The study was approved by the HCB Ethics Committee (number of approval: HCB/2021/0007). Being an observational retrospective study, written informed consent for study inclusion and consent for publication were obtained from all patients that were still alive and in follow-up at our Institution.

## Contributors

F.S. and A.P. conceived the study. T.P., M.B., F.S. and B.C. retrieved Institutional patients’ data. F.S. performed the statistical and bioinformatic analyses. F.S., F.B.M., I.D.S. and S.N. wrote the first manuscript draft. All authors contributed to the final interpretation of study data. All authors contributed to manuscript revisions and approved its final version.

## Data Availability

The data generated in this study are not publicly available due to information that could compromise patient privacy or consent but are available upon reasonable request from the corresponding author.

## Acknowledgements

F.S. is supported by a Rio Hortega clinical scientist contract from the Instituto de Salud Carlos III (ISCIII) and was supported by an ESMO Fellowship – Translational during the conduction of this study. A.P. received funding from Fundación CRIS contra el cáncer PR_EX_2021-14, Agència de Gestó d’Ajuts Universitaris i de Recerca 2021 SGR 01156, Fundación Fero BECA ONCOXXI21, Instituto de Salud Carlos III PI22/01017, Asociación Cáncer de Mama Metastásico IV Premios M. Chiara Giorgetti, Breast Cancer Research Foundation BCRF-23-198, and RESCUER, funded by European Union’s Horizon 2020 Research and Innovation Programme under Grant Agreement No. 847912. F.B-M. received funding from Fundación científica AECC Ayudas Investigador AECC 2021 (INVES21943BRAS). B.C. received funding the European Union’s Horizon 2020 research and innovation programme under the Marie Skłodowska–Curie Grant Agreement No. 955951. S.V.S. received the support of the Departament de Recerca i Universitats de la Generalitat de Catalunya (Grant No. 2021-SGR-01332, to the Molecular Image in Nuclear Medicine Research Group). M.B. is supported by a Rio Hortega clinical scientist contract from the Instituto de Salud Carlos III (ISCIII). Any views, opinions, findings, conclusions, or recommendations expressed in this material are those solely of the authors and do not necessarily reflect those of funding entities. Preliminary results of this study were presented at ESMO Breast Congress 2023 as poster presentation n. 55P^38^.

## Code availability statement

No proprietary R codes were used for the purpose of this study and all codes are retrievable online for free.

