## Supplementary methods for "Unraveling the clinicopathological and molecular changes induced by neoadjuvant chemotherapy and endocrine therapy in hormone receptor-positive/HER2-low and HER2-0 breast cancer"

*Baseline variables compared between HER2-low and HER2-0 breast tumors as secondary objective*

Age, menopausal status, estrogen receptor (ER), progesterone receptor (PgR), Ki67, stromal tumor-infiltrating lymphocytes (TILs), tumor grade, primary tumor size (cT), axillary lymph-nodes involvement (cN), histotype, PAM50 intrinsic subtype (IS), PAM50 risk-of-relapse with subtypes and proliferation (ROR-P) score and class, proportion of patients receiving neoadjuvant and adjuvant chemotherapy (CT) and endocrine therapy (ET), as well as CT/ET type.

*Study objectives*

The primary objective of this study was to compare hormone receptor-positive (HR+) HER2-0 and HER2-low cases with respect to the possible biological changes induced by the neoadjuvant strategy in terms of HER2 status, PAM50 IS and research-based PAM50 ROR-P score switch. Secondary objectives included: 1) a comparison of baseline clinicopathological and molecular features according to HER2 status; 2) a comparison of PAM50 gene expression (GE) changes induced by the neoadjuvant treatment in HER2-low vs. HER2-0 disease; 3) a comparison of surgical and survival outcomes in term of breast conservative surgery (BCS) rates, pathologic complete response (pCR), residual cancer burden (RCB), event-free survival (EFS) and overall survival (OS) according to baseline HER2 status; 4) an assessment of the association of HER2 status switch and post-surgical HER2 status with EFS and OS.

*Study endpoints definitions*

EFS was defined as the time from the first diagnosis of breast cancer until progression/relapse of the disease (local or distant) or death from any cause, whichever occurred first. OS was defined as the time from the first diagnosis of BC until the occurrence of death from any cause. RCB was originally described by Symmans et al.[1] and was hereby assessed with the MD Anderson calculator available at: http://www3.mdanderson.org/app/medcalc/index.cfm?pagename=jsconvert3, while pCR was defined as the absence of invasive tumor in-breast and in-axilla after neoadjuvant therapy and posterior surgery (i.e. ypT0/is + ypN0).

*PAM50 gene expression analysis – expanded*

RNA was purified from available archival formalin-fixed paraffin-embedded (FFPE) tumor tissues from pre-treatment baseline diagnostic biopsies and surgical specimens. RNA was extracted using the High Pure FFPET RNA isolation kit (Roche, Indianapolis, IN, USA) following the manufacturer's protocol. One to five 10-μm FFPE slides, depending on tumor cellularity, were used for each tumor sample and macrodissection performed, when needed, to avoid normal breast tissue contamination, as much as possible. A minimum of 100 ng of total RNA was analyzed at the nCounter platform (NanoString Technologies, Seattle, WA, USA) using a customized PAM50 gene panel to measure the expression of 50 BC-related genes, 4 immune-related genes (*PDCD1*, *CD274*, *CD4* and *CD8A*), androgen receptor gene (*AR*), and 5 house- keeping genes (*ACTB*, *MRPL19*, *PSMC4*, *RPLP0*, and *SF3A1*). Gene counts were log base 2 transformed and normalized using the five housekeeping genes. All tumors were assigned to an intrinsic molecular subtype (Luminal A, Luminal B, HER2-enriched, Basal-like, and Normal-like) using the research-based PAM50 subtype predictor[2]. For each available tumor sample, the correlation coefficients of all 5 IS, the PAM50's proliferation, luminal, basal and HER2 signatures, as well as the ROR score based on subtype and proliferation (ROR-P) were assessed as described elsewhere[3,4].

*Statistical analysis – expanded*

Mann-Whitney U test for unpaired data and Wilcoxon signed rank test for paired data, along with $\chi^2$ test were used to compare continuous and categorical variables between HER2-0 and HER2-low cases, respectively. PAM50 IS, ROR-P and HER2 status changes were tested with the Bhapkar or McNemar test, when appropriate, and agreement of pre/post-surgical HER2 status was assessed with Cohen's Kappa. The association of clinicopathological and molecular features with EFS and OS was assessed with Cox regression models to estimate hazard ratios (HRs) with their 95% confidence intervals (CIs). Survival curves were estimated by the Kaplan-Meier method and differences were assessed with the log-rank test. Patients alive were censored at the date of the last follow-up. Associations with BCS, RCB and pCR were conducted with logistic regressions to estimate odds ratios (ORs) with their respective 95% CIs. Significance was set at $p \leq 0.05$. Due to the exploratory nature of this study, p-values were not corrected. Two-class, paired or unpaired, significance analysis of microarrays (SAM) were used to assess GE differences between different timepoints or groups of interest, respectively.

A false discovery rate (FDR)≤5% was considered for significance. All analyses were conducted with R vers. 3.6.1[5] and SPSS® Statistics vers. 24 (IBM®, Armonk, NYC, USA) for MacOSX.
