## Supplementary table 1 for "Unraveling the clinicopathological and molecular changes induced by neoadjuvant chemotherapy and endocrine therapy in hormone receptor-positive/HER2-low and HER2-0 breast cancer"

### **Supplementary table 1. Treatments and surgical outcomes**

| **TREATMENTS AND PATHOLOGIC OUTCOMES** | **HER2-0** | | **HER2-Low** | | ***P*** |
| --- | --- | --- | --- | --- | --- |
|  | **N** | **%** | **N** | **%** |  |
|  | 69 | 37.1 | 117 | 62.9 |  |
| **NAT Type** |  |  |  |  |  |
| NACT | 40 | 58.0 | 57 | 48.7 | 0.222 |
| NET | 29 | 42.0 | 60 | 51.3 |  |
| **Neoadjuvant CT Type** |  |  |  |  |  |
| Anthracyclines + Taxanes | 39 | 97.5 | 56 | 98.2 | 0.345 |
| Taxane-based | 0 | 0.0 | 1 | 1.8 |  |
| Anthracycline-based | 1 | 2.5 | 0 | 0.0 |  |
| **Neoadjuvant ET Type** |  |  |  |  |  |
| Tamoxifen +/- GnRHa | 10 | 34.5 | 10 | 16.7 | 0.059 |
| Aromatase Inhibitor | 19 | 65.5 | 50 | 83.3 |  |
| **Type of surgery** |  |  |  |  |  |
| Conservative | 33 | 47.8 | 62 | 53.0 | 0.693 |
| Mastectomy | 36 | 52.2 | 55 | 47.0 |  |
| **Surgical management of the axilla** |  | | | | |
| SLNB | 39 | 56.5 | 69 | 58.9 | 0.687 |
| ALND | 30 | 43.5 | 47 | 40.2 |  |
| None | 0 | 0.0 | 1 | 0.9 |  |
| **Adjuvant CT** |  |  |  |  |  |
| Yes | 13 | 18.8 | 15 | 12.8 | 0.267 |
| No | 56 | 81.2 | 102 | 87.2 |  |
| **Adjuvant ET** |  |  |  |  |  |
| Yes | 67 | 97.1 | 110 | 94.0 | 0.344 |
| No | 2 | 2.9 | 7 | 6.0 |  |
| **Adjuvant RT** |  |  |  |  |  |
| Yes | 51 | 73.9 | 87 | 74.4 | 0.946 |
| No | 18 | 26.1 | 30 | 25.6 |  |
| **pCR (ypT0/is + ypN0/ITC)** |  |  |  |  |  |
| Yes | 7 | 10.1 | 14 | 12.0 | 0.705 |
| No | 62 | 89.9 | 103 | 88.0 |  |
| **RCB** |  |  |  |  |  |
| 0 | 7 | 10.1 | 14 | 12.0 | 0.740 |
| I | 12 | 17.4 | 16 | 13.7 |  |
| II | 34 | 49.3 | 65 | 55.6 |  |
| III | 16 | 23.2 | 22 | 18.8 |  |

**Legend.** NAT: neoadjuvant therapy; NACT: neoadjuvant chemotherapy; NET: neoadjuvant endocrine therapy; pCR: pathologic complete response; RCB: residual cancer burden; CT: chemotherapy; ET: endocrine therapy; GnRHa: gonadotropin-releasing hormone analogue; SLNB: sentinel lymph-node biopsy; ALND: axillary lymph-node dissection.
