## Supplementary table 2 for "Unraveling the clinicopathological and molecular changes induced by neoadjuvant chemotherapy and endocrine therapy in hormone receptor-positive/HER2-low and HER2-0 breast cancer"

**Supplementary table 2. PAM50 ROR-P and intrinsic subtypes distribution at baseline and after neoadjuvant therapy in paired samples**

| MOLECULAR FEATURES | BASELINE | | | | *P* |
|---|---|---|---|---|---|
| | HER2 0 | | HER2-Low | | |
| **Intrinsic Subtype** | *N* | *%* | *N* | *%* | |
| *Overall* | 45 | 100.0 | 78 | 100.0 | |
| Luminal A | 19 | 42.2 | 44 | 56.4 | |
| Luminal B | 16 | 35.6 | 26 | 33.3 | 0.217 |
| HER2E | 4 | 8.9 | 5 | 6.4 | |
| Basal-like | 2 | 4.4 | 2 | 2.6 | |
| Normal-like | 4 | 8.9 | 1 | 1.3 | |
| **ROR-P Category** | *N* | *%* | *N* | *%* | |
| *Overall* | 42 | 100.0 | 66 | 100.0 | |
| Low | 10 | 23.8 | 14 | 21.2 | *0.034* |
| Intermediate | 19 | 45.2 | 44 | 66.7 | |
| High | 13 | 31.0 | 8 | 12.1 | |
| Median | 40.8 | - | 32.1 | - | 0.241 |
| IQR | 13.1 - 55.0 | - | 16.0 - 44.3 | - | |
| **MOLECULAR FEATURES** | **POST-SURGERY** | | | | ***P*** |
| | **HER2 0** | | **HER2-Low** | | |
| **Intrinsic Subtype** | *N* | *%* | *N* | *%* | |
| *Overall* | 45 | 100.0 | 78 | 100.0 | |
| Luminal A | 15 | 33.3 | 41 | 52.6 | |
| Luminal B | 0 | 0.0 | 2 | 2.6 | 0.094 |
| HER2E | 0 | 0.0 | 2 | 2.6 | |
| Basal-like | 1 | 2.2 | 1 | 1.3 | |
| Normal-like | 29 | 64.4 | 32 | 41.0 | |
| **ROR-P Category** | *N* | *%* | *N* | *%* | |
| *Overall* | 42 | 100.0 | 66 | 100.0 | |
| Low | 32 | 76.2 | 52 | 78.8 | 0.272 |
| Intermediate | 10 | 23.8 | 11 | 16.7 | |
| High | 0 | 0.0 | 3 | 4.5 | |
| Median | -2.0 | - | -3.5 | - | 0.933 |
| IQR | -7.7 - 9.2 | - | -7.8 - 6.9 | - | |

**Legend.** ROR-P: risk of relapse score based on subtypes and proliferation; HER2-E: HER2-enriched; IQR: interquartile range. Significant p values in italics.
