## Supplementary table 3 for "Unraveling the clinicopathological and molecular changes induced by neoadjuvant chemotherapy and endocrine therapy in hormone receptor-positive/HER2-low and HER2-0 breast cancer"

**Supplementary table 3. PAM50 ROR-P and intrinsic subtypes distribution at baseline and after NACT and NET in paired samples according to HER2 status**

| Molecular class | HER2-0 (paired samples) | | | | *P values for subtype and ROR switch** |
|---|---|---|---|---|---|
| | *Pre NACT* | | *Post NACT* | | |
| **ROR-P** | | | | | |
| Low risk | 2 | 8.7 | 18 | 78.3 | |
| Intermediate risk | 12 | 52.2 | 5 | 21.7 | *<0.001* |
| High risk | 9 | 39.1 | 0 | 0.0 | |
| *Overall* | 23 | 100.0 | 23 | 100.0 | |
| **PAM50 IS** | | | | | |
| Luminal A | 8 | 32.0 | 7 | 28.0 | |
| Luminal B | 12 | 48.0 | 0 | 0.0 | |
| HER2-E | 3 | 12.0 | 0 | 0.0 | *<0.001* |
| Basal-like | 2 | 8.0 | 1 | 4.0 | |
| Normal-like | 0 | 0.0 | 17 | 68.0 | |
| *Overall* | 25 | 100.0 | 25 | 100.0 | |
| | *Pre NET* | | *Post NET* | | |
| **ROR-P** | | | | | |
| Low risk | 8 | 42.1 | 14 | 73.7 | |
| Intermediate risk | 7 | 36.8 | 5 | 26.3 | *0.009* |
| High risk | 4 | 21.1 | 0 | 0.0 | |
| *Overall* | 19 | 100.0 | 19 | 100.0 | |
| **PAM50 IS** | | | | | |
| Luminal A | 11 | 55.0 | 8 | 40.0 | |
| Luminal B | 4 | 20.0 | 0 | 0.0 | |
| HER2-E | 1 | 5.0 | 0 | 0.0 | *<0.001* |
| Basal-like | 0 | 0.0 | 0 | 0.0 | |
| Normal-like | 4 | 20.0 | 12 | 60.0 | |
| *Overall* | 20 | 100.0 | 20 | 100.0 | |
| **Molecular class** | **HER2-Low (paired samples)** | | | | |
| | *Pre NACT* | | *Post NACT* | | |
| **ROR-P** | | | | | |
| Low risk | 2 | 7.7 | 19 | 73.1 | |
| Intermediate risk | 18 | 69.2 | 4 | 15.4 | *<0.001* |
| High risk | 6 | 23.1 | 3 | 11.5 | |
| *Overall* | 26 | 100.0 | 26 | 100.0 | |
| **PAM50 IS** | | | | | |
| Luminal A | 12 | 37.5 | 12 | 37.5 | |
| Luminal B | 15 | 46.9 | 2 | 6.3 | |
| HER2-E | 3 | 9.4 | 1 | 3.1 | *<0.001* |
| Basal-like | 2 | 6.3 | 1 | 3.1 | |
| Normal-like | 0 | 0.0 | 16 | 50.0 | |

| | | | | | |
|---|---|---|---|---|---|
| *Overall* | 32 | 100.0 | 32 | 100.0 | |
| | *Pre NET* | | *Post NET* | | |
| **ROR-P** | | | | | |
| Low risk | 12 | 30.0 | 33 | 82.5 | |
| Intermediate risk | 26 | 65.0 | 7 | 17.5 | *<0.001* |
| High risk | 2 | 5.0 | 0 | 0.0 | |
| *Overall* | 40 | 100.0 | 40 | 100.0 | |
| **PAM50 IS** | | | | | |
| Luminal A | 32 | 69.6 | 29 | 63.0 | |
| Luminal B | 11 | 23.9 | 0 | 0.0 | |
| HER2-E | 2 | 4.3 | 1 | 2.2 | *<0.001* |
| Basal-like | 0 | 0.0 | 0 | 0.0 | |
| Normal-like | 1 | 2.2 | 16 | 34.8 | |
| *Overall* | 46 | 100.0 | 46 | 100.0 | |

**Legend.** NACT: neoadjuvant chemotherapy; NET: neoadjuvant endocrine therapy; HER2-E: HER2-enriched;. *: referred to Bhapkar tests carried out to test for subtype and ROR-P class switches. Significant p
