## Supplementary table 4 for "Unraveling the clinicopathological and molecular changes induced by neoadjuvant chemotherapy and endocrine therapy in hormone receptor-positive/HER2-low and HER2-0 breast cancer"

**Supplementary table 4. Association of HER2 status and main clinicopathological features at baseline with pathologic responses**

| Variables | pCR | | | | | | | |
|---|---|---|---|---|---|---|---|---|
| | Univariate OR | Lower 95%CI | Upper 95%CI | *P* | Multivariate OR | Lower 95%CI | Upper 95%CI | *P* |
| *ER% (continuous)* | 0.98 | 0.96 | 1.00 | *0.046* | 1.00 | 0.98 | 1.02 | 0.972 |
| *PgR% (continuous)* | 0.99 | 0.97 | 1.00 | *0.027* | 0.99 | 0.98 | 1.00 | 0.174 |
| *Ki67% (continuous)* | 1.04 | 1.02 | 1.06 | *0.001* | 1.03 | 1.00 | 1.06 | 0.101 |
| *NAT (NACT vs. NET)* | 0.15 | 0.04 | 0.54 | *0.004* | 4.48 | 1.16 | 17.35 | *0.030* |
| *TNM (II-III vs. I)* | 0.79 | 0.31 | 2.02 | 0.624 | 0.65 | 0.23 | 1.84 | 0.419 |
| *HER2 status (HER2-low vs. HER2-0)* | 1.20 | 0.46 | 3.15 | 0.704 | 1.67 | 0.57 | 4.87 | 0.345 |
| **Variables** | **RCB-0/I** | | | | | | | |
| | Univariate OR | Lower 95%CI | Upper 95%CI | *P* | Multivariate OR | Lower 95%CI | Upper 95%CI | *P* |
| *ER% (continuous)* | 0.99 | 0.97 | 1.00 | 0.067 | 1.00 | 0.98 | 1.01 | 0.582 |
| *PgR% (continuous)* | 0.99 | 0.99 | 1.00 | 0.172 | 1.00 | 0.99 | 1.01 | 0.505 |
| *Ki67% (continuous)* | 1.02 | 1.01 | 1.04 | *0.009* | 1.01 | 0.98 | 1.03 | 0.657 |
| *NAT (NACT vs. NET)* | 0.25 | 0.12 | 0.53 | *<0.001* | 4.00 | 1.72 | 9.34 | *0.001* |
| *TNM (II-III vs. I)* | 0.57 | 0.29 | 1.12 | 0.101 | 0.42 | 0.20 | 0.90 | *0.025* |
| *HER2 status (HER2-low vs. HER2-0)* | 0.91 | 0.46 | 1.78 | 0.777 | 1.04 | 0.50 | 2.16 | 0.924 |

**Legend.** ER: estrogen receptor; PgR: progesterone receptor; NAT: neoadjuvant therapy; NACT: neoadjuvant chemotherapy; NET: neoadjuvant endocrine therapy; OR: odds ratio; CI: confidence interval; pCR: pathologic complete response; RCB: residual cancer burden. Significant p values in italics.
