## Supplementary table 5 for "Unraveling the clinicopathological and molecular changes induced by neoadjuvant chemotherapy and endocrine therapy in hormone receptor-positive/HER2-low and HER2-0 breast cancer"

### Supplementary table 5. Multivariable analyses for EFS and OS

| Variables | EFS | | | |
|---|---|---|---|---|
| | HR | Lower 95%CI | Upper 95%CI | *P* |
| *Age (continuous)* | 1.04 | 0.99 | 1.09 | 0.161 |
| *cT (T3-4 vs- T1-2)* | 6.95 | 2.00 | 21.77 | *0.002* |
| *cN (positive vs. negative)* | 3.34 | 1.11 | 10.04 | *0.032* |
| *PgR% (continuous)* | 0.97 | 0.96 | 0.99 | *0.005* |
| *Ki67% (continuous)* | 1.01 | 0.98 | 1.04 | 0.725 |
| *HER2 shift (stable vs. changed)* | 550393.13 | 3.13E-135 | 9.69E+148 | 0.936 |
| *CT treated (yes vs. no)* | 0.73 | 0.20 | 2.69 | 0.631 |
| **Variables** | **OS** | | | |
| | **HR** | **Lower 95%CI** | **Upper 95%CI** | ***P*** |
| *Age (continuous)* | 1.08 | 1.01 | 1.16 | *0.030* |
| *cT (T3-4 vs- T1-2)* | 20.71 | 2.93 | 146.42 | *0.002* |
| *cN (positive vs. negative)* | 11.74 | 1.28 | 107.51 | *0.029* |
| *PgR% (continuous)* | 0.97 | 0.94 | 1.00 | 0.074 |
| *Ki67% (continuous)* | 1.02 | 0.98 | 1.05 | 0.368 |
| *HER2 shift (stable vs. changed)* | 806319.14 | 2.1768E-179 | 2.987E+193 | 0.950 |
| *CT treated (yes vs. no)* | 0.51 | 0.08 | 3.36 | 0.487 |

**Legend.** cT: baseline clinical primary tumor dimension; cN: baseline clinical axillary nodal involvement; CT: chemotherapy; HR: hazard ratio; CI: confidence interval; PgR: progesterone receptor. Significant p values in italics.
