## Supplementary figure 2 for "Unraveling the clinicopathological and molecular changes induced by neoadjuvant chemotherapy and endocrine therapy in hormone receptor-positive/HER2-low and HER2-0 breast cancer"

**Supplementary figure 2. DGE at baseline and after surgery between HER2-Low and HER2-0 overall and according to neoadjuvant treatment strategy**

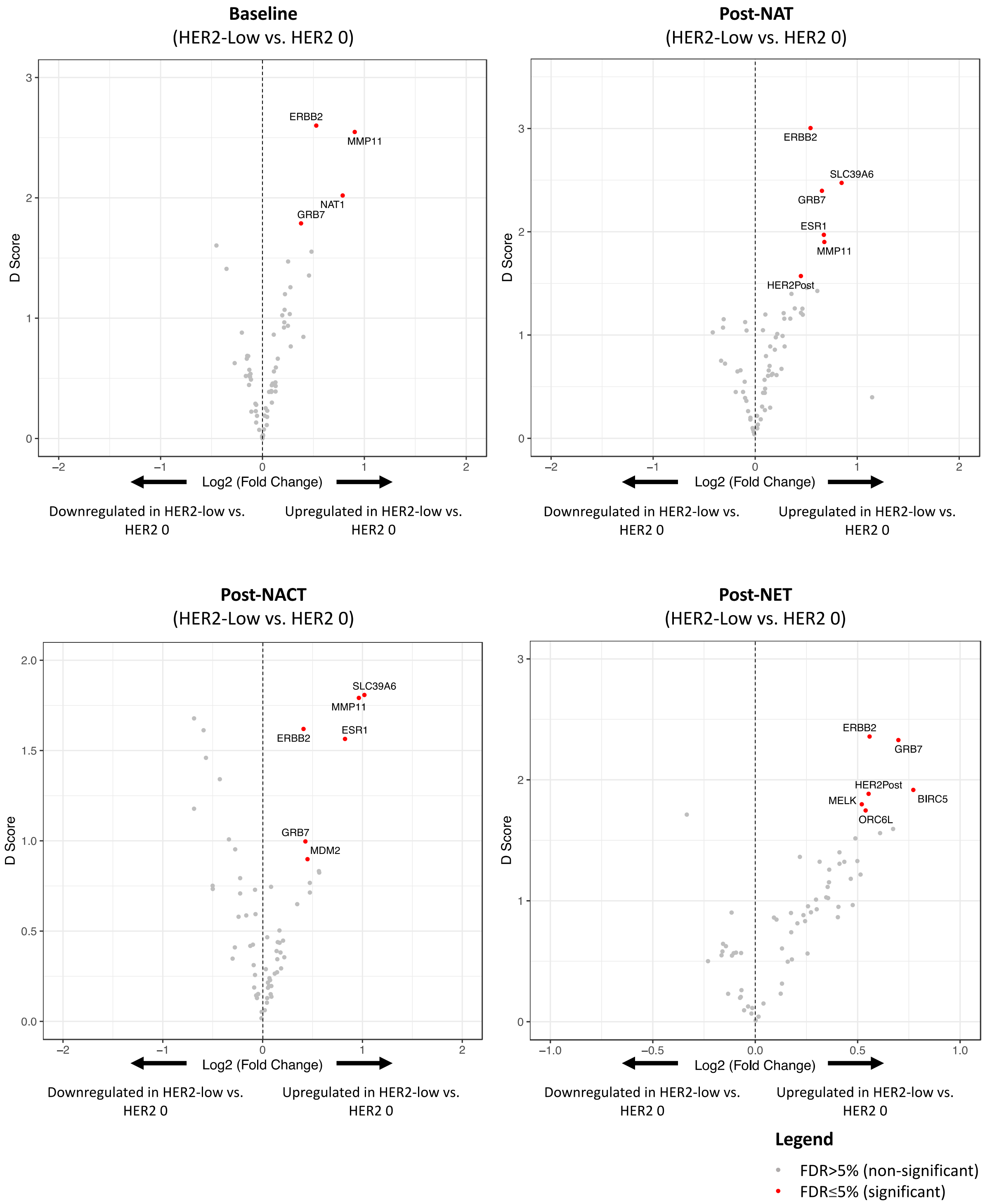


**Legend.** DGE: differential gene expression; NAT: neoadjuvant therapy; NET: neoadjuvant therapy; NACT: neoadjuvant chemotherapy; FDR: false discovery rate; D score: a T-statistic value that reflects a standardized change in expression. It measures the strength of the relationship between gene expression and the HER2-low category (versus HER2-0); Grey dots represent genes not differentially expressed, while red dots identify significantly differentially expressed genes.
