## Supplementary figure 1 for "Unraveling the clinicopathological and molecular changes induced by neoadjuvant chemotherapy and endocrine therapy in hormone receptor-positive/HER2-low and HER2-0 breast cancer"

### Supplementary figure 1. STROBE flow-chart

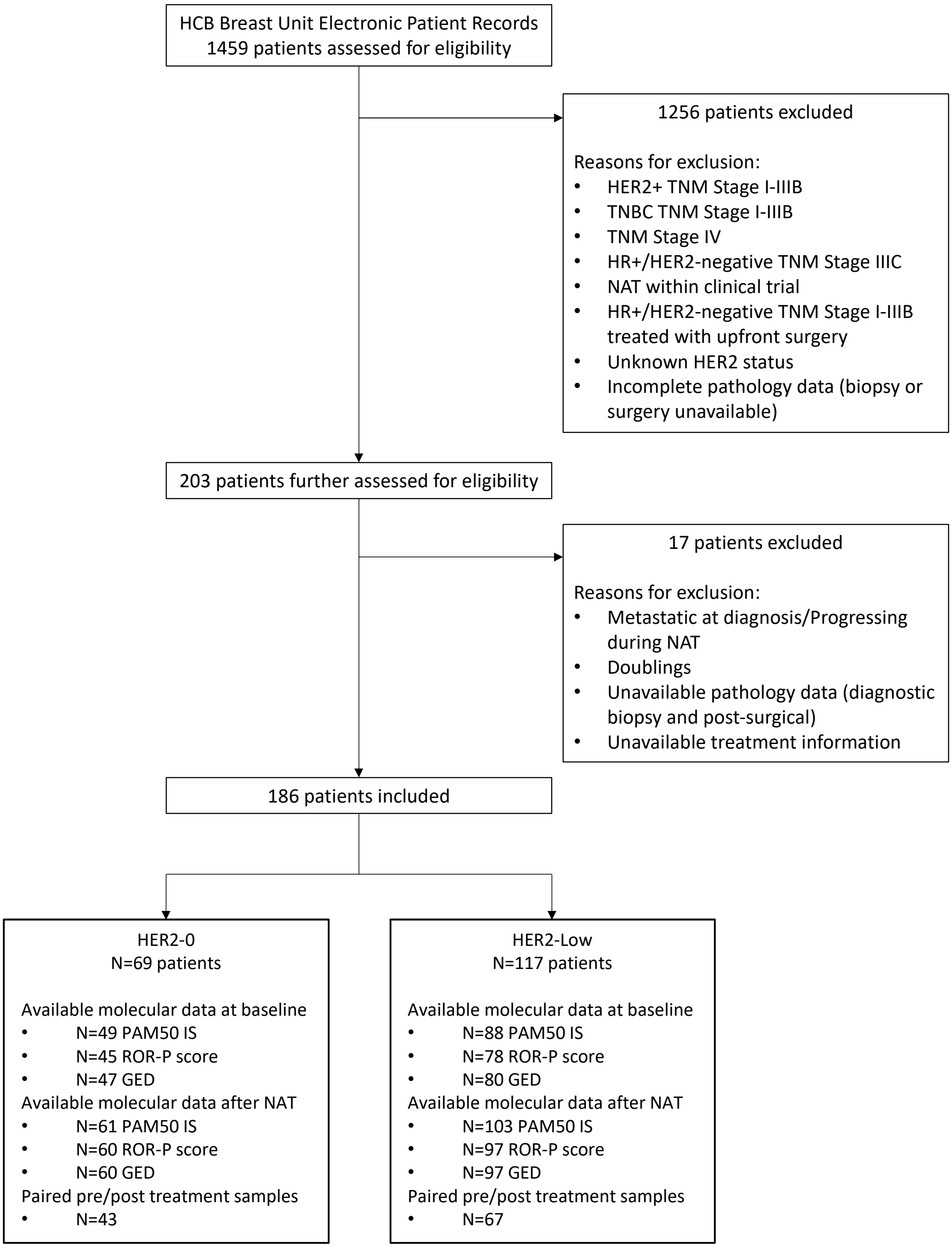


**Legend.** GED: gene expression data; ROR: risk of relapse score; IS: intrinsic subtypes; HCB: Hospital Clinic of Barcelona; NAT: neoadjuvant therapy; TNBC: triple negative breast cancer; +: positive; HR: hormone receptors.
